# Can a herd immunity strategy become a viable option against COVID-19? A model-based analysis on social acceptability and feasibility

**DOI:** 10.1101/2020.05.19.20107524

**Authors:** Takashi Akamatsu, Takeshi Nagae, Minoru Osawa, Koki Satsukawa, Takara Sakai, Daijiro Mizutani

## Abstract

This paper studies the social acceptability and feasibility of “herd immunity” strategies against coronavirus disease 2019 (COVID-19). To this end, we propose a control scheme that aims to develop herd immunity while satisfying the following two basic requirements for a viable policy option. The first requirement is social acceptability: the overall deaths should be minimized for social acceptance. The second is feasibility: the healthcare system should not be overwhelmed to avoid diverse adverse effects. Exploiting the fact that the severity of the disease increases considerably with age, the proposed control scheme protects high-risk individuals that mainly consist of the elderly. The protection of high-risk individuals reduces the average severity of the disease in the unprotected population, thereby substantially reducing mortality and avoiding the collapse of the healthcare system. Social acceptability (in terms of the resulting mortality) and feasibility (in terms of the healthcare system capacity) of the proposed herd immunity strategy are summarized into two respective, easily computable conditions by building on a simple susceptible–infected–recovered (SIR) model. For its parsimony, the proposed framework can provide a rule of thumb that applies to various populations for studying the viability of herd immunity strategies against COVID-19. For Japan, herd immunity may be developed by the considered control scheme if 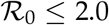 and the severity rates of the disease are 1/10 times smaller than the previously reported value, although as high mortality as seasonal influenza is expected.

## 1 Introduction

The pandemic of coronavirus disease 2019 (COVID-19) has been a huge global health threat, with over 4.5 million cases and 300, 000 deaths confirmed worldwide as of the 17th May 2020 [1]. It has been observed from experience from China, Italy, and the United States that COVID-19 can overwhelm the healthcare capacities. Due to the absence of an effective antiviral drug or a vaccine, many countries have adopted non-pharmaceutical interventions (NPIs), such as closing schools and workplaces and imposing rigorous social distancing measures to reduce transmission of the virus. Initial efforts by governments worldwide have been concentrated on the short-run *suppression* of the first wave of the pandemic to avoid the collapse of the healthcare system. Suppression of an outbreak can, however, leave a large portion of the population uninfected and susceptible; therefore, the possibility exists for a resurgence of outbreaks as severe as the initial one [2, 3, 4]. Moreover, radical NPIs aimed at suppression are not sustainable, as they have already had devastating impacts on the global economy and personal lives of many. Long-run strategies that go beyond simple relaxation of the initial suppression-oriented responses are crucial, as we probably are at least one or two years away from the substantial supply of a vaccine. The continued circulation of the virus in the global population for a prolonged period seems to be inevitable. Then, a possible option might be developing *herd immunity* by ensuring a sufficiently high proportion of immune individuals in the population, which may be achieved through a carefully managed slow spread of infection that avoid the collapse of the healthcare system [5].

Social acceptance is a significant issue for any form of herd immunity policies because they are associated with considerable numbers of infections and hence mortality. For an infectious disease with a basic reproduction number 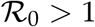, herd immunity requires that a proportion 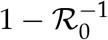 of the whole population must be infected at least. For COVID-19, the basic reproduction number had been reported to be around 2.5 [6, 7, 8], whereas the infection fatality ratio (IFR) around 0.6 to 0.7% [9, 10, 11]. A crude estimate by these numbers is that the resulting mortality along the way toward herd immunity can reach 400 per 100, 000, or about 500, 000 deaths for Japan and 1.3 million deaths for the United States. These numbers would not be socially acceptable. The basic premise of our research is that the true IFR for COVID-19 is lower than the reported value since no herd immunity approaches can obtain social acceptance otherwise.

We propose a long-run control strategy that aims at developing herd immunity and study its social acceptability (in terms of the resulting mortality) and feasibility (in terms of the healthcare capacity). Although uncertainties remain for COVID-19, age is a key factor for the severity of the disease, with fatality increasing disproportionately with age. For instance, those with age ≥ 80 can be 1, 000 to 10, 000 times more likely to die upon infection than those with < 20 according to previous estimates [9, 11]. By exploiting this fact, the proposed control scheme consists of two components. The first component is a protection measure for high-risk individuals (i.e., the elderly and those with underlying health conditions) to reduce mortality. The second component is a dynamic control measure (time-dependent reductions of social activities) for the unprotected to avoid the collapse of the healthcare system. The protection of high-risk individuals reduces the average severity ratio of the disease for the unprotected population and has twofold implications. First, it contributes to social acceptance because it can substantially reduce mortality. Second, it contributes to feasibility because severe cases remain below the healthcare system’s capacity even when infections spread in the unprotected, low-risk population. With these effects, the number of infected individuals can increase without causing a collapse of the healthcare system. Our analyses reveal that, if several conditions are met with the basic reproduction number of the epidemic, 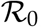, and several other characteristic parameters of the system, the whole population can acquire herd immunity without the collapse of the healthcare system, while substantially reducing mortality.

Rather than simulating highly structured models, we build on the basic susceptible-infected-recovered (SIR) model [12]. At the cost of the simplification, we can study the feasibility and social acceptability of herd immunity strategies semi-analytically with a limited number of input variables. The only required inputs are the age composition of the population, the healthcare system’s service capacity, and the age-specific severity rates for COVID-19 while we acknowledge that uncertainties remain for the last one. For its simplicity, the proposed framework is universally applicable to different populations without resorting to data-intensive and model-specific structural simulations and calibrations. For Japan, as an example, herd immunity can be developed if 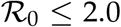 and the severity rates of the disease (including the IFR) is 1/10 times smaller than the previously reported value [9]. Such numerical results obtained by the proposed framework can be easily updated by incorporating the latest estimates for the input parameters.

## 2 The control scheme

A population of a fixed size faces with a novel infectious disease with a basic reproduction number 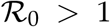. We propose a control scheme that consists of the two components discussed below. See Appendix A.1 for the mathematical details.

(a) **Protection measure for high-risk individuals**. To minimize fatality from an outbreak, some forms of protection for high-risk individuals must be in place [13]. Existing evidence consistently suggests that age is a key factor that affects the severity of COVID-19, with fatality ratio substantially increasing for the elderly [9, 11]. To this end, the whole population is partitioned into two groups: the *protected* and the *active*. The former group consists of high-risk individuals, such as the elderly or those with pre-existing medical conditions, who are likely to die if they become infected. The latter consists of low-risk individuals who can recover from infection with high probability. The former group is safely isolated from the active group and protected from the disease during the outbreak. Individuals in the latter group can interact with others in the same group and thus may be infected. The size of the active group is normalized to unity. The size of the whole population, including both the protected and the active, is denoted by *N* ≥ 1. The relative size of the active group in the whole population, *n* ≡ 1/*N* ∊ (0, 1], is chosen in the first place.

**Figure 1:**
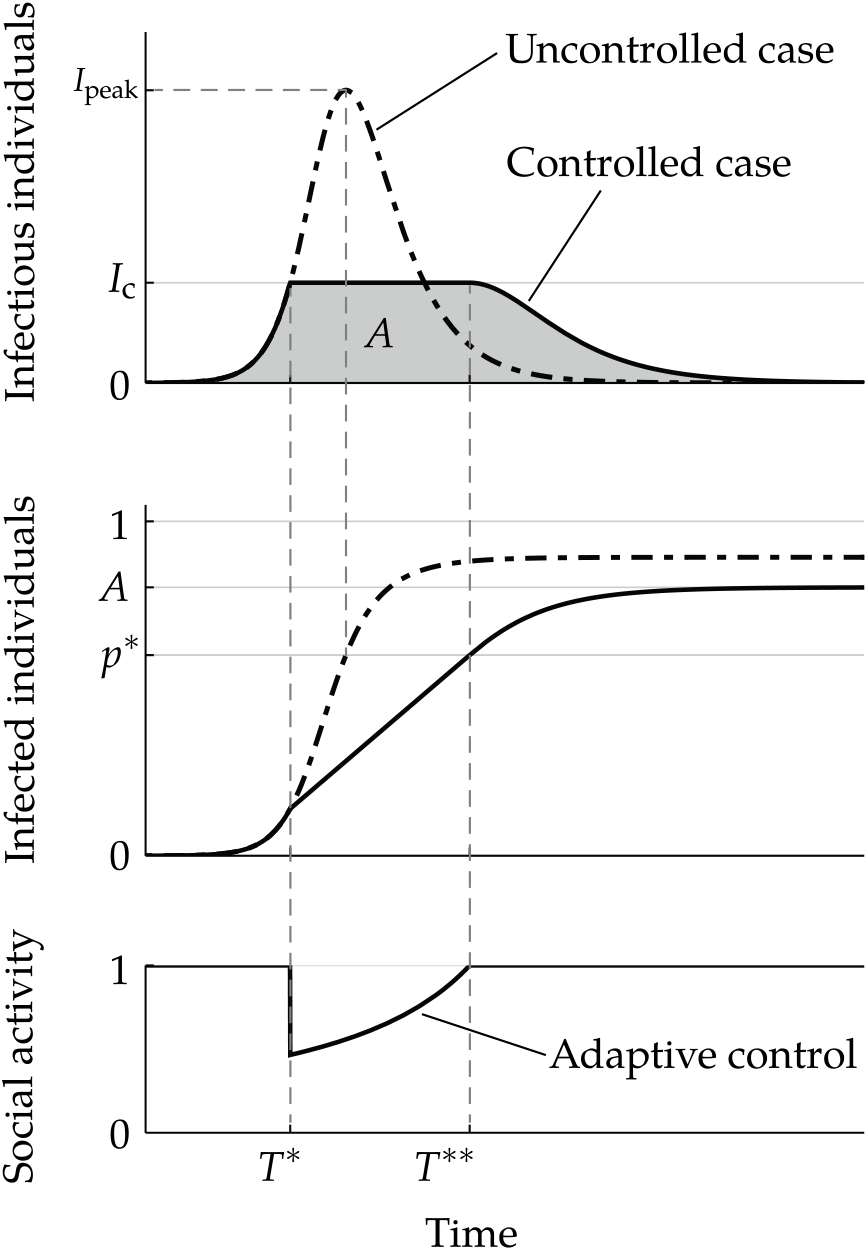
Illustration of the proposed adaptive control measure. The top and middle shows, respectively, the number of infectious individuals *I*(*t*) and infected (infectious or recovered) individuals 1 − *S*(*t*) at time t. The uncontrolled case is shown by the dot-dashed curve, whereas a controlled case by the black solid curve. An adaptive control that reduces social activity as in the bottom panel is imposed at *T\** and lifted when herd immunity in this group is established at *T*\*\*. The area of the gray region below the controlled curve in the top panel is the final number of infected individuals in the active group, A. Since the number of infectious individuals start to decrease at *T*\*\*, *A* will exceed 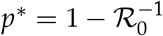.

(b) **Adaptive control measure for low-risk individuals** (see Figure 1). As the disease can spread within the active group, some dynamic control measures should be imposed on this group to avoid the collapse of the healthcare system. Let *I*(*t*) be the number of infectious individuals in the active group at time *t*. To keep *I*(*t*) below the maximum level that the healthcare system can handle, the proposed control scheme has a predetermined threshold *I_c_* for *I*(*t*) and aims to maintain

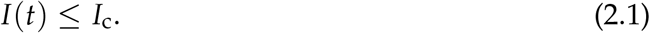

To achieve this, after the onset of the outbreak in the active group, the social activity level at t is set to

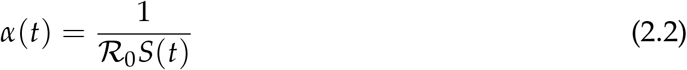

once *I*(*t*) reaches *I_c_* at some time *T*\*, as shown in the bottom panel of Figure 1. By this adaptive control, the effective reproduction number *α*(*t*)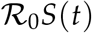 is kept at 1, so that *I*(*t*) stays at the threshold level *I_c_* during the peak period (the top panel of Figure 1). At some time *T*\*\* > *T*\*, the remaining number of susceptible individuals equals 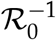, or the cumulative number of infected individuals equals

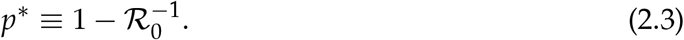

The proportion *p*\*∊ (0, 1) is the well-known *herd immunity threshold*, or the threshold share of infected individuals for a population to develop herd immunity. Thus, herd immunity *for the active group* (not necessarily for the whole population, including the protected) is developed at *T*\*\*. Without any control, the number of infectious individuals in the active group declines after *T*\*\* due to herd immunity. When the outbreak within the active group goes to an end, the protection of high-risk individuals can be lifted. The area below the epidemic curve in the top panel of Figure 1 corresponds to the final number of recovered individuals, which we denote by A.

## 3 Social acceptability and feasibility

There are three conditions under which the proposed control policy can be a viable policy option against a novel infectious disease such as COVID-19. First, for social acceptance, the overall mortality should not become too large. Second, the healthcare system should not be overwhelmed. Third, the final number of infected individuals, A, should be sufficiently high to acquire herd immunity. The last two conditions may be called the feasibility conditions. If the proposed strategy is both socially acceptable and feasible, herd immunity approaches of the proposed type would be viable policy options.

### Social acceptability: Upper bound for the overall mortality

The first component (a) of the control scheme aims at reducing mortality. Suppose that we can accept only 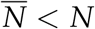 lost lives due to the epidemic. Then, the following condition must be met:

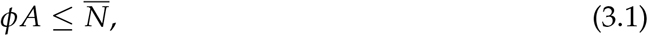

where *ϕ* ∊ (0, 1) is the average mortality rate in the active group upon infection and A the final cumulative number of infections in the active group. The left-hand side of (3.1) is the number of lives lost by the end of the outbreak. We say the proposed policy is (*socially*) *acceptable* if condition (3.1) is met.

The condition (3.1) restricts the size of the active group relative to the whole population. The larger the active group, the more high-risk individuals are included in it. Thus, *ϕ* increases with the size of the active group. Evidence suggests that the fatality ratio for the elderly is by far higher than that for the young [9]. The flip side of this is that we can substantially reduce the average mortality rate *ϕ* for the active group by imposing the protection measure (a), thereby relaxing (3.1) without the difficult compromise of increasing 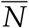 in the first place.

### Feasibility (1): Healthcare system’s capacity

The second component (b) of the proposed control scheme aims at acquiring herd immunity with limited healthcare resources. The first feasibility requirement for this component is that the control threshold *I_c_* must be lower than the *effective capacity* of the healthcare system, which we denote by *I*_max_, for the instantaneous number of infectious individuals. For instance, let *μ >* 0 denote the per capita beds available for the care of severe cases that require hospitalization, i.e., the *service capacity* of the healthcare system. To avoid various adverse effects, including a surge in mortality, the service capacity must not be exceeded. Suppose that a given proportion, *θ* ∊ (0, 1), of infected individuals in the active group needs hospitalization. Then, for the healthcare system to maintain its normal functioning, we must require *θ*I_c_ *≤ Nμ*, that is,

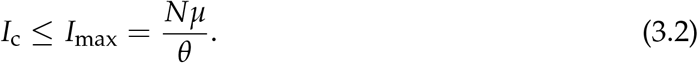

Thus, *I*_max_ = *Nμ*/*θ* gives the effective capacity. If *θ* stands for the proportion of infected individuals who need critical care, then the service capacity *μ* should be replaced with the per capita intensive care unit (ICU) beds.

The effective capacity *I*_max_ is decreasing in *θ* and thus is minimized when no protection measure for high-risk individuals is in place. Equivalently, *I*_max_ is increasing in *N* because *θ* is decreasing in *N*. Also, *I*_max_ is increasing in *N* because the per capita service capacity *for the active group* is *N_μ_*. Therefore, by protecting the elderly and other high-risk individuals, we can considerably increase *I*_max_ without enhancing the service capacity *μ*.

### Feasibility (2): Acquisition of herd immunity

The last condition requires that the final number of infected individuals in the active group, *A*, must be sufficiently high to acquire herd immunity for the whole population. Specifically, *A* should satisfy the following condition for herd immunity *as the whole population* to be acquired:

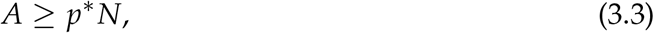

where we recall that 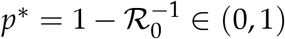 is the herd immunity threshold. If (3.3) is met, the protection measure for high-risk individuals can be lifted after a sufficient drop in the number of infected individuals in the active group without causing a secondary outbreak.

**Figure 2:**
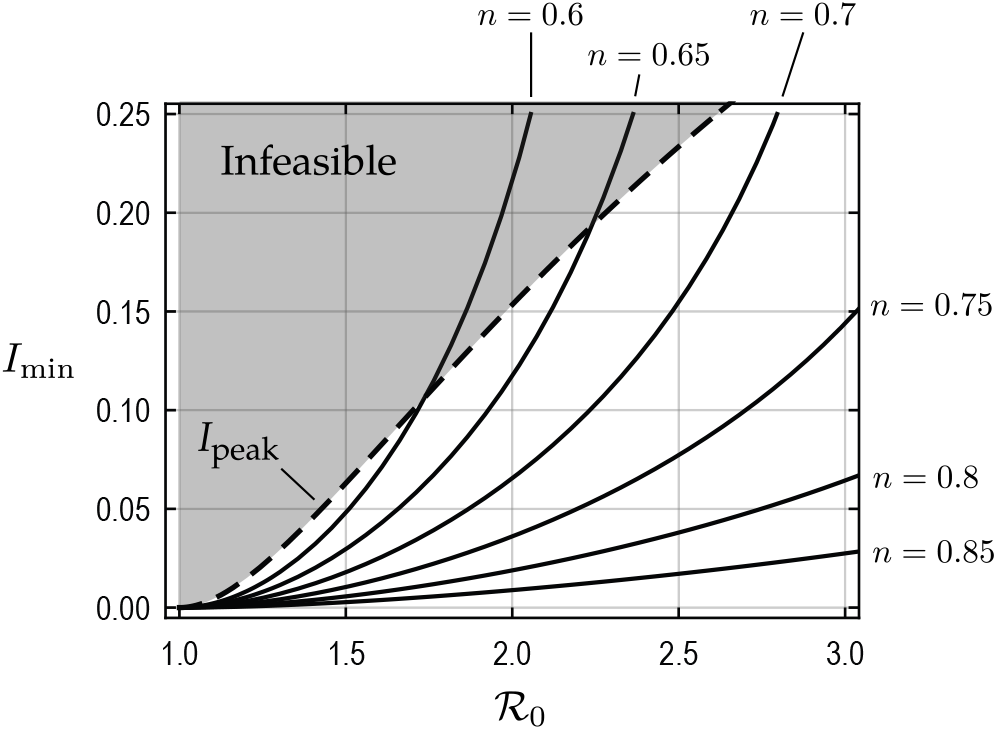
Minimum level of the control threshold to acquire herd immunity with varying shares of the active group in the whole population, *n* = 1/*N* ∊ (0, 1]. For each *n*, the condition (3.3) is satisfied if *I_c_* ≥ *I*_min_. The proposed strategy with any control threshold, *I_c_*, that is above the black solid curve can achieve herd immunity as the whole population. The dashed curve indicates the peak number of infectious individuals *I*_peak_, which is an increasing function of 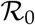, in the uncontrolled scenario. The assumed control scheme is feasible (in terms of the condition (3.3)) if *I*_min_ ≤ *I*_peak_.

From the middle panel of Figure 1, we see that *A* is an increasing function of *I_c_* so long as *I*_c_ ≤ *I*_peak_. Then, because *A* must be sufficiently high, there is a minimum value *I*_min_ for *I*_c_ so that the whole population can acquire herd immunity if *I*_c_ ≥ *I*_min_. The threshold, *I*_min_, is an increasing function of 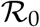 and *N*, as the right-hand side of (3.3) is increasing in both 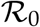 and *N*. See (A.8) in Appendix A.1 for the concrete formula of *I*_min_. Figure 2 shows the minimum level *I*_min_ against 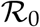 for different choices of *n* = 1/ *N* ∊ (0, 1]. Because *I*_c_ cannot exceed the peak level for the uncontrolled scenario, the assumed control scheme is feasible only if *I*_min_ ≤ *I*_peak_ (see Figure 1).

By the two conditions (3.2) and (3.3), the proposed control requires *I*_c_ to satisfy *I*_min_ ≤ *I*_c_ ≤ *I*_max_ and *I*_c_ ≤ *I*_peak_. For this reason, it must be that We say the proposed strategy is *feasible* if (3.4) is met.

**Table 1:**
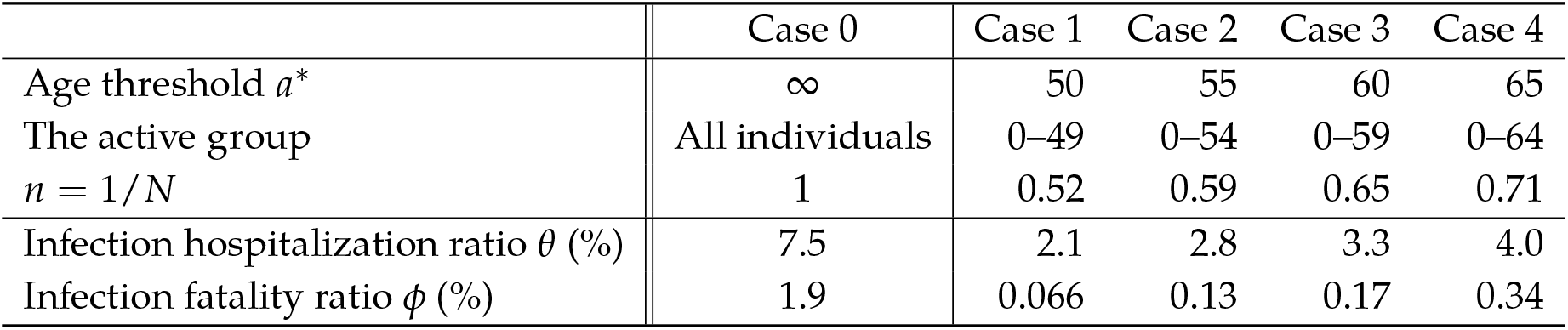
Infection hospitalization ratio and infection fatality ratio for the active group with different *a*\* under the Japanese population composition. The age-specific severity ratios are adopted from [9], whereas the Japanese population data from [14]. Case 0 (no protection measure) is shown for reference.

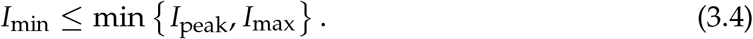

### Summary of the variables

The variables that appear in our framework are as follows. The policy variables that characterize the proposed control scheme are the size of the active group *n =* 1/*N* and the control threshold *I*_c_. The environmental constants related to the disease, which cannot be modified by the policy, are 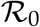, *θ*, and *ϕ*, where the latter two are increasing functions of n. The other constants are the per capita healthcare service capacity *μ*, which can be increased in the long run, and the socially acceptable mortality 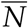.

## 4 Numerical illustration with Japan

Taking Japan as an example, this section illustrates how we can study the social acceptability and feasibility of the proposed control scheme. With age being a key factor for the severity of COVID-19, a pragmatic way to define the active group would be choosing a threshold age *a*\*, whereby those individuals with age < *a*\* constitute the active group. To obtain *θ* and *ϕ* numerically as functions of *a*\*, we simply combine an available estimate for the age-specific infection hospitalization ratio (IHR) and infection fatality ratio (IHR) for COVID-19 ([9], Tables 1 and 3) and the latest estimate for the population composition of the country [14] (see Appendix A.2).

Table 1 summarizes the basic parameters *n*, *θ*, and *ϕ* for the active group with different choices of *a*\*. Case 0 is the uncontrolled scenario without any protection or dynamic control measures. Case 1, 2, 3, and 4 correspond, respectively, to the proposed control scheme with *a*^*^ = 50, 55, 60, and 65. As seen, both infection hospitalization ratio (IHR) *θ* and infection fatality ratio (IFR) *ϕ* increases with *a*\*. For all Cases 1 to 4, both IHR and IFR are substantially lower than Case 0 (no protection). Below, we examine the social acceptability and feasibility conditions discussed in Section 2 using the baseline parameters shown in the table.

### Social acceptability

We observe that the proposed approach, or herd immunity strategies in general, would not become socially acceptable for Japan if the reported estimate of the IFR for COVID-19 is not an overestimation. Table 2 shows the minimum mortality that results from the proposed herd immunity strategy in Cases 1 to 4 under different values of 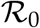. The minimum mortality is obtained simply by multiplying the Japanese population by *ϕp\**, where 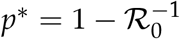 is the herd immunity threshold. The condition (3.1) requires that the resulting mortality should not become too large. Table 2 clearly indicates that herd immunity approaches are not socially acceptable if the reported IFR is close to the “true” value. For instance, 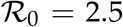 must result in at least 130,000 deaths in Case 3. Although it is around 1/10 times smaller than the uncontrolled scenario with 1.4 million deaths (Case 0), such a number would not be socially acceptable for a country with less than 1,000 confirmed COVID-19 deaths (as of 15 May 2020). In particular, Case 4 would not become socially acceptable for any 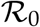 shown in the table.

**Table 2:** The minimum mortality to acquire herd immunity (thousand deaths). A blank cell indicates that the share of the active group *n* for that settings is too small to develop herd immunity for the whole population (i.e., *n* < *p*\*).

| $\mathcal{R}_0$ | $p^*$ | Case 0 | Case 1 | Case 2 | Case 3 | Case 4 |
| --- | --- | --- | --- | --- | --- | --- |
| 1.5 | 0.41 | 790 | 27 | 53 | 72 | 140 |
| 2.0 | 0.50 | 1,200 | 41 | 80 | 110 | 210 |
| 2.5 | 0.60 | 1,400 |  |  | 130 | 260 |

That said, if the reported IFR is true, *any form of* herd immunity strategies would not be socially acceptable for the country. It can be the case that, however, the adopted baseline estimate of the IFR is a pessimistic bound. For instance, various reports suggest that there is a large number of asymptomatic infections [15, 16, 17], which can imply *θ* and *ϕ* are smaller than the baseline estimates. For Japan, a recent study based on serological testing argues that there might have been around 400-to 850-fold infections more than confirmed cases with PCR testing in Kobe City, Japan [18]. This result can be an overestimation, with its all limitations, including the specificity of the employed test kit and selection bias. On the other hand, these studies consistently suggest that true IFR can be much smaller than the previous estimates due to asymptomatic or mild infections.

Below, as a thought experiment, we assume that the IFR *ϕ* (and also the IHR *θ* for consistency) is 1/10 times smaller than the reported estimate by [9]. Under this assumption, the resulting mortality due to the proposed strategy becomes the order of 10, 000 for Japan (Case 3), which is akin to average annual influenza-related mortality in the country.

### Feasibility

Next, we study feasibility taking Case 3 (*a*\* *=* 60) as the example. Based on a report from the Ministry of Health, Labor and Welfare, we assume that the hospital beds count available for COVID-19 patients in Japan is 3.1 × 10^4^, or *μ =* 2.5 × 10^−4^ per capita [19]. The effective capacity is then given by

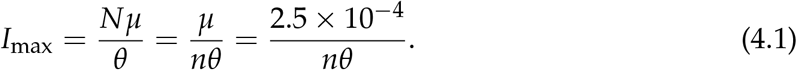

The effective capacity becomes a similar value even if we use the proportion of infections that require critical care for *θ* and replace *μ* with the per capita free ICU beds (see Appendix A.2). Table 3 shows the values of the effective capacity for all Cases 0 to 4, where *θ* is assumed to be 1/10 times the adopted IHR [9] to ensure consistency with the corresponding assumption on *ϕ*. We see that the protection measure substantially increases *I*_max_ in all Cases 1 to 4 compared with Case 0. For instance, in Case 3, *I*_max_ is around 3.4 times greater than Case 0.

**Figure 3:**
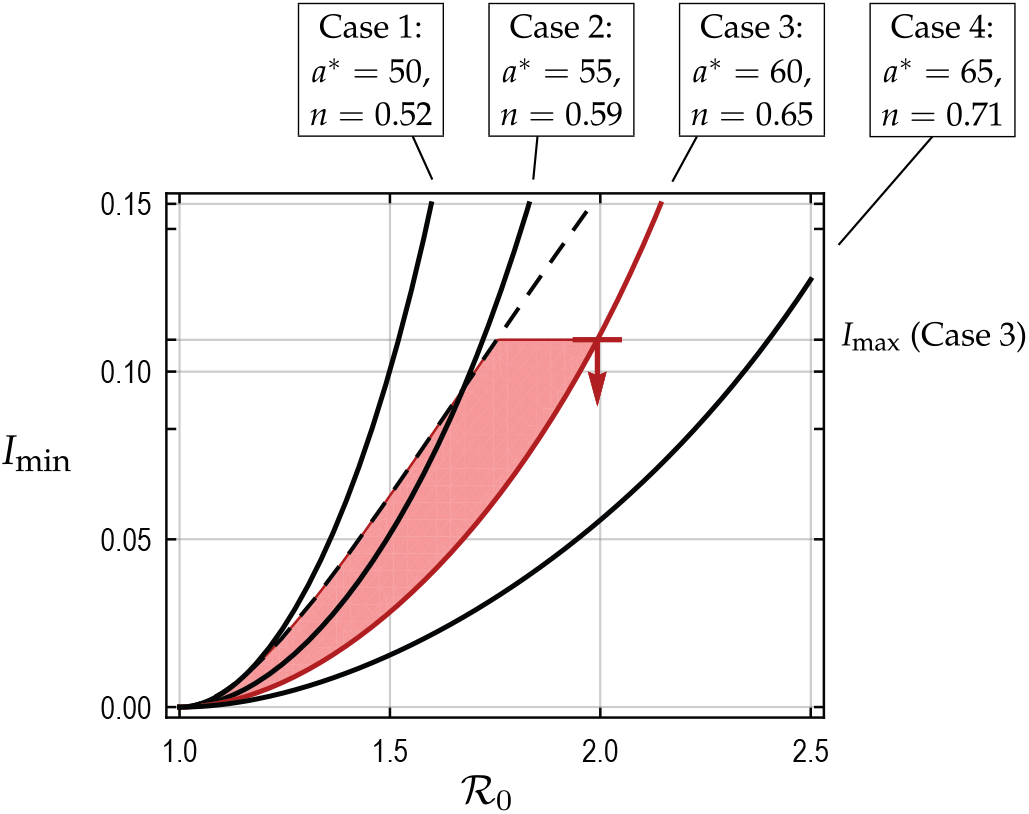
Feasibility of the proposed control scheme for Japan. The solid black curves show *I*_min_ for Cases 1 to 4. The dashed black curve shows *I*_peak_. The marker indicates *I*_max_ for Case 3, below which the proposed control scheme is feasible in terms of the healthcare system capacity. In Case 3, the proposed control scheme is feasible when 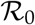 and *I*_c_ lie in the shaded region. The feasibility condition (3.4) is thus satisfied for 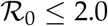 in Case 3.

**Table 3.** Effective healthcare capacity *I*_max_.

|  | Case 0 | Case 1 | Case 2 | Case 3 | Case 4 |
| --- | --- | --- | --- | --- | --- |
| $I_{\max}$ | 0.0330 | 0.221 | 0.147 | 0.113 | 0.0857 |

Figure 3 examines the feasibility of the proposed control scheme for Case 3. The solid black lines indicate *I*_min_ for different *a\**, which are obtained by using the corresponding shares *n* of the active group (Table 1). Cases 1, 2, and 4 are also shown for comparison. The dashed curve indicates *I*_peak_ as in Figure 2, while the marker on *I*_min_ in Case 3 shows *I*_max_ for that case. The condition (3.4) means that the proposed control scheme is feasible if *I*_min_ stays below both *I*_peak_ and *I*_max_. For Case 3, *I*_max_ = 0.113 and feasibility in terms of the healthcare capacity is satisfied for 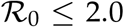. As we see *I*_max_ ≤ *I*_peak_ when 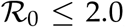, Case 3 satisfies the condition (3.4), and hence is feasible, if 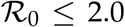. If 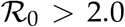 is the case, a substantial increase in the hospital beds would in order unless *θ* is smaller than our assumption.

## 5 Discussions

This paper provides a simple framework for studying the viability of herd immunity strategies as policy options against COVID-19. We propose a control strategy with a protection measure for the elderly and other high-risk individuals. By the protection measure, the proposed control scheme aims at (i) minimizing the resulting mortality along the way toward herd immunity and (ii) preventing the collapse of the healthcare system by reducing the proportion of severe cases during the outbreak. The thought experiment with Japanese parameters suggests that the proposed strategy can be an acceptable and feasible option if the “true” severity of the disease (i.e., the IHR *θ* and the IFR *ϕ*) is lower than the reported value by [9]. In concrete terms, if the severity of the disease is 1/10 times lower than the reported estimate, the proposed control scheme can become a viable policy option when 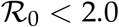.

Apart from our numerical examples, which can be easily updated and corrected using the latest data, a more important qualitative observation is that the protection measure for high-risk individuals is the key to the social acceptability and feasibility of herd immunity approaches. For instance, even if the final number of infected individuals is the same, the resulting mortality is reduced considerably by imposing a protection measure (Table 2). Likewise, without increasing the baseline service capacity (hospital beds), the effective capacity of the healthcare system Imax increases significantly by a protection measure (Table 3). These results stem from the fact that the severity of COVID-19 increases disproportionately with age. For this reason, it is expected that relatively young populations may be able to acquire herd immunity without causing too many deaths per capita, as argued in [20]. Also, it is observed that, whether we aim at herd immunity or not, some protection measures for high-risk individuals can substantially relax healthcare capacity constraints.

### Related literature

The proposed control scheme reduces the resulting mortality by minimizing the “overshoot” from the herd immunity threshold for the whole population (in the notation of Section 2, we aim at *A* = *p*\**N*), as discussed in [2]. The dynamic control measure considered in this paper, which primarily aims to develop herd immunity, is conceptually different from previous studies that focus on suppression in the short run. For instance, some studies consider intermittent lock-down strategies that aim to keep the number of critically ill patients below the ICU service capacity [3, 21]. Because simple suppression policies do not protect high-risk individuals and thus must face with a considerably smaller effective capacity (compare Case 0 with, e.g., Case 3 in Table 3). Therefore, even if herd immunity might be developed by intermittent suppression without a protection measure for high-risk individuals as in [21], the duration of such a control strategy would become by far longer than that with protection, thereby placing a substantial burden on the economy, not to mention the resulting mortality.

### Limitations

Our framework has several apparent limitations, of which we highlight the following two.

First, our mathematical model is a stylized simplification of any reality. We assume the simplest SIR model as the epidemic dynamics, which may overestimate the herd immunity threshold *p*\* [22]. For simplicity, we consider a strict protection measure for high-risk individuals under which they are entirely confined, but a more realistic approach would be age-diversified social distancing measure as considered in [13, 23]. Reliable estimates for the number of infected individuals in the population are crucial for implementing an adaptive control scheme as the proposed one and assessing how close it is to herd immunity. The introduction of systematic antibody tests at scale would be of high priority [5, 24, 25].

It will also provide better estimates for the severity of the disease. Second, our analyses assume lifelong immunity after recovery for simplicity. However, how long immunity against severe acute respiratory syndrome-coronavirus 2 (SARS-CoV-2) lasts, or whether we can develop sufficient immunity against SARS-CoV-2 in the first place, are still unknown. If sufficient immunity against the virus can not be developed, herd immunity strategies, in general, would fail. Due to unanswered questions about the pathogen, including the ones mentioned above, extreme care should be taken before adopting any form of herd immunity policies. Should a herd immunity policy be adopted despite the expected risks, our analysis suggests that the protection measure of high-risk individuals is a promising way to grant both social acceptability and feasibility.

## Data Availability

All the data used in the manuscript are publicly available.

## A Technical appendix

### A.1 The epidemic model

We consider the basic SIR model [12]. A single population of a fixed size faces with a novel infectious disease. There are two groups of agents, the protected and the active. The former is safely isolated from the virus in the first place. The total mass of individuals in the active group is normalized to unity. The relative size of the whole population is *N ≥* 1, thereby 1/*N* ∊ (0,1] is the share of the active group in the population. Below, *S*, *I*, and *U* denote, respectively, susceptible, infectious, and recovered individuals, where we have *S* + *I* + *U* = 1. We use *U* for recovered individuals to avoid confusion with reproduction numbers. The dependence of these quantities on time *t* is suppressed where no confusion should result.

#### Uncontrolled dynamics

The uncontrolled dynamics of the epidemic in the active group are given by the following equations: *Ṡ =* − *βSI*, *İ* = *βSI* − *γI*, and 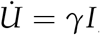, where *β >* 0 is the infection rate and γ *>* 0 the removal rate of infectious individuals, which are unchangeable constants. The initial condition is (*S*(0), *I*(0), *U*(0)) = (1 − *ϵ*, *ϵ*,0) where ϵ *>* 0 is a tiny number. Under the dynamics, 1/*γ* is the average duration of infection (say, 20 to 30 days for COVID-19, including the incubation period).

We scale time by the unit of the average duration of infection 1/*γ*, so that the dynamics reduce into the following dimensionless equations: 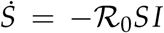, 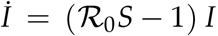, and 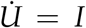, where 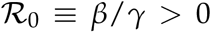 is the *basic reproduction number*. We assume 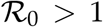, for which an outbreak occurs from any small *I*(0) > 0 if there are no control measures. By using this time scale, the area below the epidemic curve {*I*(*t*) | *t* > 0} coincides with the final number of infected individuals.

The quantity 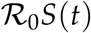 is the *effective reproduction number*, which represents transmissibility at time *t*, for the uncontrolled dynamics. If 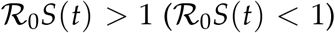, then *I*(*t*) increases (decreases) in *t*. This implies that, for the uncontrolled case, there is a threshold *S*\* for *S*(*t*),

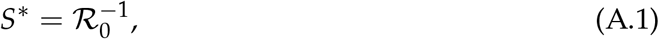

and *I*(*t*) is maximized at *t*\* such that *S*(*t*\*) = *S*\*; the threshold proportion of the cumulative number of infectious individuals to develop herd immunity, or the *herd immunity threshold*, is given by 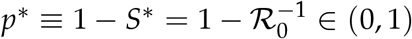. The peak number of infectious individuals in the uncontrolled case is given by

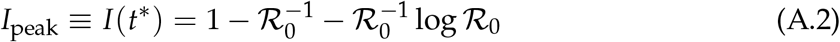

since 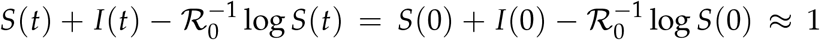 for all *t*. As is well known, the final number of infected individuals 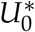 for the uncontrolled scenario is the unique solution for 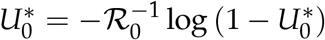.

#### Controlled dynamics

We assume that we have full control over the social activity level in the active group so that only a time-dependent fraction *α*(*t*) ∊ [0,1] of active individuals can contact with others. The controlled epidemic dynamics become 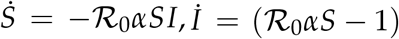, and 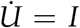. The effective reproduction number for the controlled dynamics is given by 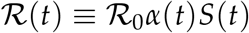. With a control threshold *I*_c_, the proposed adaptive control is the following (see the bottom panel of Figure 1):

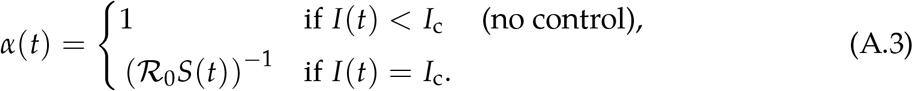

Under the above control, 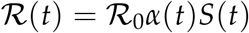 satisfies

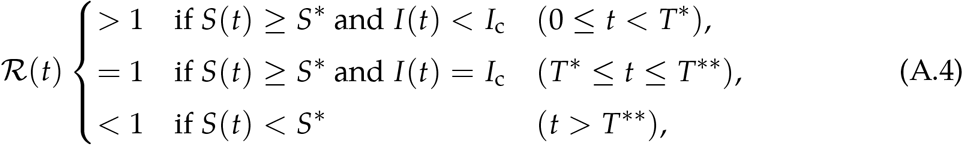

where *T*\* is the initial time the number of infectious individuals *I*(*t*) reaches *I*_c_ and *T*\*\* is the time the active group acquires herd immunity, i.e., *S*(*T*\*\*) = *S*\* (see Figure 1). After *T*\*\*, the outbreak in the active group goes to an end without any control. Standard final size arguments give the final size of the epidemic in the active group. Under the assumed SIR model, we have the identity

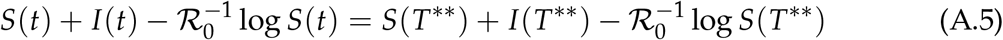

at any *t* after *T*\*\*. The initial conditions at 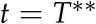 are 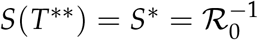 and 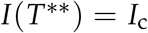, provided that *I*_c_ ≤ *I*_peak_, whereas the terminal conditions for *t* → ∞ are *I*(∞) − 0 and 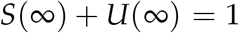. Plugging these into (A.5), the *final size* 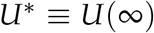 of the epidemic in the active group under the proposed control policy is given as the solution for the following equation:

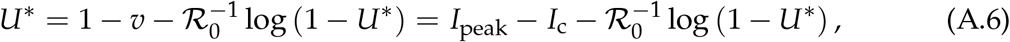

where 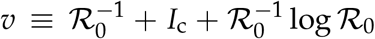 is the value of the right hand side of (A.5). By the implicit function theorem regarding (A.6), we observe

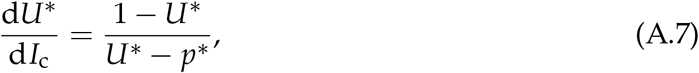

implying that 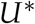 is increasing in *I*_c_ because 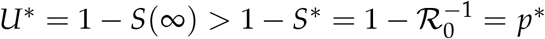.

**Figure 4:**
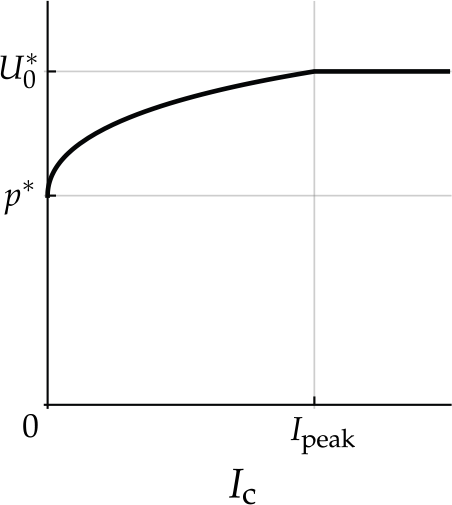
The final number of infected individuals 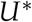 as a function of *I*_c_ under a given 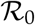. That in the uncontrolled case is denoted by 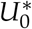. The minimum level for *I*_c_, *I*_min_, is the solution for 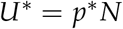, which exists only if 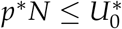.

The protection of high-risk individuals can be lifted at some *T*\*\*\* ≫ *T* ** where *I* (*T*\*\*\*) is sufficiently small so that 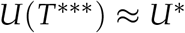. After the protection measure is lifted at time *T*\*\*\*, the size of the population becomes *N*. Then, no second outbreak can occur if 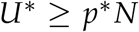, which implies (3.3) where we denote 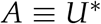.

Plugging the minimum final size 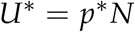 into (A.6), we obtain

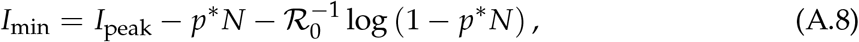

provided that 1 − *p*\**N* > 0, or *n* ≡ 1/*N* > *p*\*.

The above derivations for *I*_min_ assume that *I*_c_ ≤ *I*_peak_. If *I*_min_ > *I*_peak_ happens, it indicates the proposed control is infeasible because we cannot increase 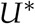 by increasing *I*_c_ beyond the uncontrolled peak level *I*_peak_ (see the top panel Figure 1). Figure 4 shows the final size 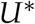 as a function of *I*_c_, which indicates that 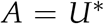 is an increasing function of *I*_c_ so long as *I*_c_ ≤ *I*_peak_. We also have 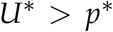 for any *I*_c_ > 0. However, 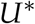 can not be increased beyond the uncontrolled case 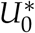.

### A.2 Data

#### Computation of *θ* and *ϕ*

We adopt the age-specific infection hospitalization ratio (IHR) and infection mortality ratio (IFR) provided in, respectively, Table 3 and 1 of [9]. Another estimates for the IFR and IHR can be found in [11], but yield similar results. We weight-average these ratios by the population composition of Japan [14] to obtain *d* and *$* as (discrete) functions of the threshold age *a*\* ∊ {5,10,15,…,55,60,65,…}, up to the availability of the population composition data. Figure 5 shows the computed profiles of *θ* and *ϕ* against *a*\* for Japan. Those for Great Britain are shown as reference, where the population composition data for Great Britain is obtained from [26]. For both countries, *θ* and *ϕ* are increasing in *a*\*. Due to Japan's aging population, the curves for the country are steeper than those for Great Britain. We note that the adopted IFRs by [9] was obtained by correcting the case fatality ratios (CFR) for under-ascertainment, which roughly corresponds to the halving of the CFR for Wuhan data, or an ascertainment ratio of 0.5. Our assumption in Section 4 of 1/10 times smaller IFR/IHR can thus loosely be interpreted as the assumption that there are 20 times more infections than confirmed cases.

**Figure 5:**
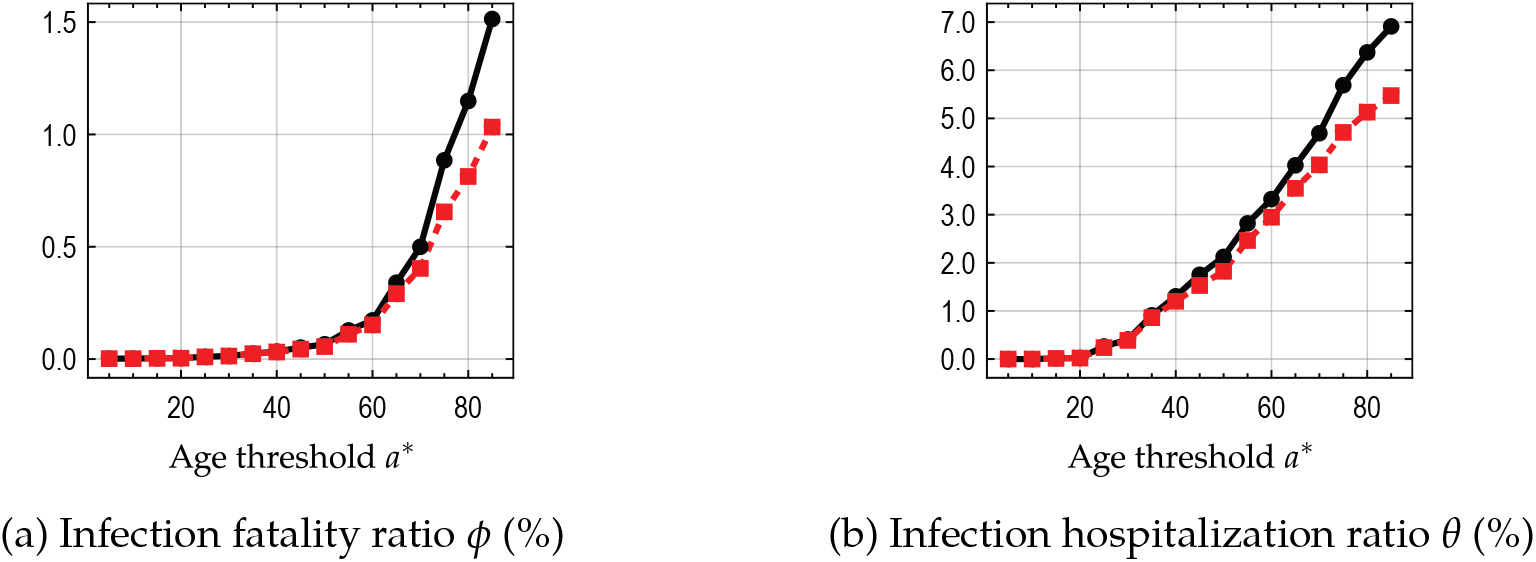
Infection hospitalization ratio *θ* and infection fatality ratio *ϕ* for the active group with different age thresholds *a*\*. The active group consists of those with age < *a*\*. Black solid curve indicates Japan, whereas red dashed curve shows Great Britain as a reference. As Japan has more aging population than Great Britain, both *θ* and *ϕ* for the country rise faster with *a*\* than those for Great Britain.

#### Healthcare service capacity *μ*

The baseline par capita service capacity for Japanese healthcare system is set to be *μ* = 3.1 × 10^4^/1.26 × 10^4^ = 2.5 × 10^−4^ following a report by the Ministry of Health, Labor and Welfare, which states that the country will prepare 31,077 hospital beds dedicated to COVID-19 [19].

#### Feasibility in terms of the critical care service capacity

Results become similar when we consider an effective capacity based on the ICU service capacity instead of hospitalization. The proportion of individuals who need critical care, which we denote by *θ′*, is reported to be around 10~20% among those hospitalized [6, 11]. As a conservative assumption, we let *θ*′ = 0.2 × *θ* where *θ* is the IHR. The total ICU and high care unit (HCU) beds count for Japan is around 1.7 × 10^4^, or *μ′* = 1.3 × 10^−4^ per capita [27]. These numbers imply 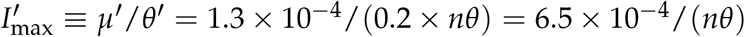, which is more than twice greater than *I*_max_ = 2.5 × 10^−4^/(*nθ*) in (4.1). However, the actual effective capacity for critical care would be somewhere close to (4.1) as we acknowledge that (i) the above 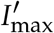 is an overestimation because the *free* ICU beds per capita must be much smaller than *μ*′ and that, on the other hand, (ii) assuming the free ICU beds in the normal setting would be an underestimation because some special reinforcement measures for COVID-19 must be in place. Because the actual situations regarding the free ICU capacity for COVID-19 are unknown, the main text uses the hospitalization capacity in the numerical illustration in Section 4.

## References

[1] World Health Organization. Coronavirus disease (COVID-2019) situation report 118, 10:00 CEST, 17 May 2020. Available from: https://www.who.int/emergencies/diseases/novel-coronavirus-2019/situation-reports/.

[2] Andreas Handel, Ira M. Longini Jr., and Rustom Antia. What is the best control strategy for multiple infectious disease outbreaks? Proceedings of the Royal Society B: Biological Sciences, 274(1611):833–837, 2007.

[3] Neil Ferguson, Daniel Laydon, Gemma Nedjati Gilani, et al. Report 9: Impact of non-pharmaceutical interventions (NPIs) to reduce COVID19 mortality and healthcare demand. Imperial College London, 16 March 2020. https://doi.org/10.25561/77482.

[4] Patrick G.T. Walker, Charles Whittaker, Oliver Watson, et al. Report 12: The global impact of covid-19 and strategies for mitigation and suppression. Imperial College London, 26 March 2020. https://doi.org/10.25561/77735.

[5] Marius Gilbert, Mathias Dewatripont, Eric Muraille, Jean-Philippe Platteau, and Michel Goldman. Preparing for a responsible lockdown exit strategy. Nature Medicine, pages 1–2, 2020.

[6] Andrea Remuzzi and Giuseppe Remuzzi. COVID-19 and Italy: What next? The Lancet, 2020.

[7] Qun Li, Xuhua Guan, Peng Wu, Xiaoye Wang, Lei Zhou, Yeqing Tong, Ruiqi Ren, Kathy SM Leung, Eric HY Lau, Jessica Y Wong, et al. Early transmission dynamics in wuhan, china, of novel coronavirus–infected pneumonia. New England Journal of Medicine, 2020.

[8] Joseph T. Wu, Kathy Leung, and Gabriel M. Leung. Now casting and forecasting the potential domestic and international spread of the 2019-nCoV outbreak originating in Wuhan, China: A modelling study. The Lancet, 395(10225):689–697, 2020.

[9] Robert Verity, Lucy C. Okell, Ilaria Dorigatti, Peter Winskill, Charles Whittaker, et al. Estimates of the severity of coronavirus disease 2019: A model-based analysis. The Lancet Infectious Diseases, 2020. https://doi.org/10.1016/S1473-3099(20)30243-7.

[10] Timothy W. Russell, Joel Hellewell, Christopher I. Jarvis, Kevin Van Zandvoort, Sam Abbott, Ruwan Ratnayake, Stefan Flasche, Rosalind M. Eggo, W. John Edmunds, and Adam J. Kucharski. Estimating the infection and case fatality ratio for coronavirus disease (COVID-19) using age-adjusted data from the outbreak on the Diamond Princess cruise ship, February 2020. Eurosurveillance, 25(12):6–10, 2020.

[11] Henrik Salje, Cécile Tran Kiem, Noémie Lefrancq, Noémie Courtejoie, Paolo Bosetti, Juliette Paireau, Alessio Andronico, Nathanaël Hozé, Jehanne Richet, Claire-Lise Dubost, Yann Le Strat, Justin Lessler, Daniel Levy-Bruhl, Arnaud Fontanet, Lulla Opatowski, Pierre-Yves Boelle, and Simon Cauchemez. Estimatingtheburden of sars-cov-2 in france. Science, 2020.

[12] William Ogilvy Kermack and Anderson G. McKendrick. A contribution to the mathematical theory of epidemics. Proceedings of the Royal Society: Series A, 115(772):700–721, 1927.

[13] Andreas Handel, Joel Miller, Yang Ge, and Isaac Chun-Hai Fung. If long-term suppression is not possible, how do we minimize mortality for COVID-19 and other emerging infectious disease outbreaks? medRxiv, 2020.

[14] Ministry of Internal Affairs and Communications. Annual report: Population estimates, 2019. https://www.e-stat.go.jp/en/stat-search/files?&toukei=%0A00200524&tstat=000000090001&stat_infid=000031921672.

[15] Hendrik Streeck, Gunther Hartmann, Martin Exner, and Matthias Schmid. Vorläufiges Ergebnis und Schlussfolgerungen der COVID-19 Case-Cluster Study (Gemeinde Gangelt), 9 April 2020. Universität-sklinikum Bonn.

[16] Kenji Mizumoto, Katsushi Kagaya, Alexander Zarebski, and Gerardo Chowell. Estimating the asymptomatic proportion of coronavirus disease 2019(COVID-19) cases on board the diamond princess cruise ship, Yokohama, Japan,2020. Eurosurveillance, 25(10):2000180, 2020.

[17] Desmond Sutton, Karin Fuchs, Mary D’Alton, and Dena Goffman. Universal screening for SARS-CoV-2 in women admitted for delivery. New England Journal of Medicine, 2020. c2009316.

[18] Asako Doi, Kentaro Iwata, Hirokazu Kuroda, Toshikazu Hasuike, Seiko Nasu, Aya Kanda, Tomomi Nagao, Hiroaki Nishioka, Keisuke Tomii, Takeshi Morimoto, and Yasuki Kihara. Estimation of sero-prevalence of novel coronavirus disease (COVID-19) using preserved serum at an outpatient setting in Kobe, Japan: A cross-sectional study. medRxiv, 2020.

[19] Labour Ministry of Health and Welfare. Number of beds for novel coronavirus infections, 2020. https://www.mhlw.go.jp/content/10900000/000628694.pdf (accessed 14 May 2020).

[20] Ari Altstedter. Infect everyone: How herd immunity could work for poor countries. Bloomberg, 21 April 2020. https://www.bloomberg.com/news/articles/2020-04-21/a-herd-immunity-strategy-could-actually-work-in-youthful-india (accessed 23April 2020).

[21] Stephen M. Kissler, Christine Tedĳanto, Edward Goldstein, Yonatan H. Grad, and Marc Lipsitch. Pro-jecting the transmission dynamics of sars-cov-2 through the postpandemic period. Science, 2020.

[22] M. Gabriela M. Gomes, Ricardo Aguas, Rodrigo M. Corder, Jessica G. King, Kate E. Langwig, Caetano Souto-Maior, Jorge Carneiro, Marcelo U. Ferreira, and Carlos Penha-Goncalves. Individual variation in susceptibility or exposureto SARS-CoV-2 lowers the herd immunity threshold. medRxiv, 2020.

[23] Daron Acemoglu, Victor Chernozhukov, Iván Werning, and Michael D Whinston. A multi-risk SIR model with optimally targeted lockdown. Technical report, National Bureau of Economic Research, 2020. NBER Working Paper No. 27102.

[24] Amy K. Winter and Sonia T. Hegde. The important role of serology for COVID-19 control. The Lancet Infectious Diseases, 21 April 2020. https://doi.org/10.1016/S1473-3099(20)30322-4.

[25] Nicholas C. Grassly, Marga Pons-salort, Edward P. K. Parker, Peter J. White, et al. Report 16: Role of testing in COVID-19 control. Imperial College London, 23 April 2020. Avail-able from: https://www.imperial.ac.uk/mrc-global-infectious-disease-analysis/covid-19/covid-19-reports/ (accessed 24 April 2020).

[26] Office for National Statistics. Estimates of the population for the UK, England and Wales, Scotland and Northern Ireland, 2020. Available from: https://www.ons.gov.uk/peoplepopulationandcommunity/populationandmigration/populationestimates/ (accessed 29 April2020).

[27] The Japanese Society of Intensive Care Medicine. Number of ICU/HCU beds by prefecture, 2020. https://www.jsicm.org/news/upload/icu_hcu_beds.pdf (accessed 15 May 2020.

